# Anatomical and Etiologic Predictors of Oculopalatal Tremor Phenotypes: A Pooled Patient-Level Analysis

**DOI:** 10.64898/2026.11.23.26368778

**Authors:** Hajar Alammar, Inaya Chowdhary, Aasef G. Shaikh

## Abstract

Oculopalatal tremor (OPT) follows disruption of the cerebello-rubro-olivary network and is frequently associated with hypertrophic inferior-olive degeneration (HOD). HOD does not invariably produce OPT; determinants of oculo-palatal, ocular-only, palatal-only, or non-oscillatory phenotypes remain undefined. We analyzed 501 patients with HOD, of which 424 with classifiable laterality: 52.1% had unilateral and 47.9% bilateral HOD. Lesion location predicted HOD laterality (chi-square=35.65, p=3.4×10^-7; Cramer V=0.31): brainstem/pons lesions produced ipsilateral HOD in 70.0%, whereas cerebellar lesions produced contralateral HOD in 86.4%. Bilateral HOD reflected combined cerebellar/brainstem lesions, and was associated with ocular oscillations (OR 2.66, 95% CI 1.31-5.43; p=0.0067) but not convincingly palatal oscillations (OR 1.49, 95% CI 0.91-2.42; p=0.137). Phenotype differed by etiology (p=0.0024): hemorrhagic and vascular-malformation lesions preferentially produced combined OPT, ischemic lesions were enriched among ocular-only presentations, and neoplastic lesions produced the most heterogeneous phenotype and were disproportionately represented among palatal-only cases. Age, sex, and lesion laterality were not associated with phenotype. Four-class prediction was limited (macro AUC 0.555; balanced accuracy 0.301), with little improvement after adding HOD laterality (macro AUC 0.563; balanced accuracy 0.323). Among classifiable OPT waveforms, 96.9% contained a pendular component. Median OPT latency was 6 months (IQR 4.75-24); lesion location, etiology, and HOD laterality did not predict latency. HOD is therefore an important substrate, but lesion anatomy and etiology shape group-level phenotype without enabling reliable individual prediction.

## Introduction

Oculopalatal tremor (OPT) is an acquired neurological syndrome characterized by rhythmic ocular oscillations accompanied by palatal tremor and most commonly arises after structural injury involving brainstem or cerebellar pathways^12^. Symptomatic palatal tremor is distinguished from essential palatal tremor by its association with structural lesions of the brainstem or cerebellum and with secondary degeneration of the inferior olive^3,4^. Progressive ataxia and palatal tremor represents an additional, clinically heterogeneous syndrome in which similar circuitry may be affected in the setting of progressive cerebellar disease^5^.

The anatomical substrate of symptomatic palatal tremor and OPT is disruption of the cerebellar-rubro-olivary system, traditionally conceptualized as the Guillain-Mollaret triangle^16^. Deep cerebellar nuclei efferents ascend through the superior cerebellar peduncle and decussate in the midbrain, whereas descending fibers involving the central tegmental tract project toward the ipsilateral inferior olive^7,8^. Interruption of these pathways produces trans-synaptic degeneration of the inferior olivary nucleus. Unlike most degenerative processes, the affected olive may enlarge, producing the characteristic radiographic appearance of hypertrophic olivary degeneration (HOD)^6, 9^. Longitudinal MRI analyses indicate that increased T2 signal can become evident approximately one month after the causative lesion, whereas frank olivary hypertrophy generally emerges later, beginning at approximately six months and potentially persisting for several years^10^.

HOD and the clinical syndrome of OPT, however, are not synonymous. Patients with apparently similar lesions may develop combined ocular and palatal oscillations, palatal oscillations without ocular involvement, ocular oscillations without palate involvement, or radiographic HOD without either manifestation^11–13^. Detailed ocular motor studies similarly demonstrate that inferior olivary abnormalities do not invariably predict the complete oculopalatal phenotype. In a series of patients with OPT-spectrum disease, ocular oscillations were predominantly vertical with variable torsional and horizontal components, several patients showed dissociated oscillations, and some patients with inferior olivary signal abnormalities never developed palatal tremor^11^.

The ocular oscillations of classical OPT are usually acquired pendular oscillations and are frequently vertical, torsional, or multidimensional^11, 14^. complex, irregular structure is difficult to explain by a single autonomous oscillator^14, 15^. Experimental and computational work has therefore proposed a dual-mechanism model^14–16^. Following loss of normal afferent input, inferior olivary neurons hypertrophy and develop increased electrotonic coupling, allowing the inferior olive to generate synchronized low-frequency oscillatory activity^1, 14^. That signal alone is insufficient to reproduce the large and irregular ocular oscillations observed clinically; maladaptive cerebellar learning is proposed to amplify and reshape the olivary output^15,14^.

This oscillator-amplifier model places OPT within a distributed olivocerebellar network rather than attributing the clinical syndrome solely to inferior olivary hypertrophy^14,2^. Contemporary reviews similarly emphasize that OPT emerges from interruption of the cerebello-rubro-olivary pathway followed by secondary olivary and cerebellar network reorganization^17^.

Despite this mechanistic framework, several clinically important questions remain unresolved. First, the relationship between the anatomical level of the primary lesion and the side of subsequent HOD has not been quantified across a large, heterogeneous patient population. Second, it remains uncertain why bilateral or unilateral HOD leads to ocular oscillations in some patients but palatal manifestations or no oscillatory syndrome in others^11,12^. Third, whether the nature of the primary lesion - hemorrhagic, ischemic, neoplastic, vascular malformation, inflammatory, traumatic, or degenerative - modifies the subsequent clinical phenotype remains incompletely established^9, 12^. Finally, OPT is recognized as a delayed neurological complication, but its typical latency and the extent to which lesion characteristics predict the timing of onset remain uncertain^7, 17, 18^.

Because OPT is uncommon, individual clinical series are generally small^11,14^. Conversely, the published literature contains hundreds of individually described patients in whom lesion location, lesion etiology, HOD characteristics, ocular manifestations, palatal manifestations, and associated neurological features can be reconstructed. We combined these observations to create a pooled patient-level dataset of 501 patients. The objectives of this study were to determine: (1) how primary lesion location and etiology relate to HOD laterality; (2) whether bilateral versus unilateral HOD, lesion location, and lesion etiology predict ocular oscillations, palatal oscillations, or combined OPT; (3) whether OPT, isolated ocular oscillation, and isolated palatal oscillation represent distinguishable clinicoradiological phenotypes; (4) the ocular and associated neurological phenomenology of these syndromes; and (5) the expected latency from the causative lesion to OPT. We hypothesized that anatomical extent and bilateral involvement of the olivocerebellar network would be more informative than right-versus-left HOD in determining the ultimate clinical phenotype.

## Methods

### Study Design and Case Ascertainment

We pooled patient records from published reports and a retrospective review of our clinical practice to examine associations among lesion characteristics, hypertrophic olivary degeneration (HOD), and ocular and palatal oscillations. The dataset comprised 501 patients, including 469 literature cases (Supplementary Methods S1) and 32 previously unpublished clinical cases. Patients were the unit of analysis. We searched PubMed from inception through June 2026 for reports of HOD, oculopalatal tremor (OPT), or related disorders of the cerebello-rubro-olivary pathway. We screened reports for extractable patient-level data and reviewed potentially overlapping reports to avoid duplicate inclusion (Supplementary Methods S2). We examined the clinical series separately for potential referral effects. The University Hospitals Cleveland Medical Center Institutional Review Board approved the retrospective chart review.

### Data Abstraction and Phenotype Classification

We used standardized abstraction to record demographics, lesion location and etiology, radiographic HOD and its laterality, ocular and palatal findings, associated neurologic manifestations, symptom latency, MRI timing, and treatment. Two reviewers independently assessed a random 30% sample of included publications for eligibility, patient identification, and consistency of clinical and imaging abstraction. Discrepancies were resolved by consensus (Supplementary Methods S3).

HOD classification followed the source imaging interpretation. For clearly lateralized lesions, classifiable HOD was coded as ipsilateral, contralateral, or bilateral relative to the lesion (Supplementary Methods S4-S5). When ocular and palatal oscillations were both explicitly documented as present or absent, we classified patients as having OPT, ocular oscillations only, palatal oscillations only, or neither. The complete phenotype cohort included 187 patients. Of these, 142 had classifiable lesion location, etiology, and HOD pattern for association analyses (Supplementary Methods S6). A separate predictive cohort included 159 patients with confirmed HOD and complete four-class phenotype assignment. Unreported findings were treated as missing, and denominators reflected available data for each analysis (Supplementary Methods S3). Ocular waveform, palatal phenomenology, and associated neurologic manifestations were characterized descriptively (Supplementary Methods S7-S8). Symptom latency was measured from the causative lesion to reported clinical onset. MRI timing was analyzed separately as the interval from lesion to imaging (Supplementary Methods S9).

### Statistical Analysis

We summarized categorical variables as counts and percentages and continuous variables primarily as medians and interquartile ranges. We assessed associations of lesion location, etiology, and HOD pattern with phenotype using omnibus Pearson chi-square tests, with Cramér V to quantify association strength. Fisher exact tests were used for pairwise or sparse comparisons. For relevant contrasts, we estimated odds ratios with 95% confidence intervals, using the group with neither ocular nor palatal oscillations as the reference for phenotype comparisons. We compared onset latency across lesion location, etiology, and HOD laterality groups using Kruskal–Wallis tests (Supplementary Methods S10).

Given sparse outcome categories, particularly the palatal-only phenotype, we used class-balanced L2-penalized multinomial logistic regression. The primary antecedent-factor model included age, sex, primary-lesion etiology, lesion location, and lesion laterality. A secondary imaging-enhanced model added HOD pattern as bilateral, left unilateral, right unilateral, or unavailable. Performance was estimated with repeated stratified four-fold cross-validation and summarized by macro one-versus-rest area under the receiver operating characteristic curve (AUC), balanced accuracy, macro F1 score, and multiclass log loss.

## Results

### Study Population, Data Completeness, Clinical Phenotype Distribution

The pooled dataset contained 501 patient records, including 469 literature-derived cases and 32 novel unpublished cases from our clinical practice. Table 1 summarizes analytical populations.

**Table 1.** Study populations and distribution of complete clinical phenotypes. Panel A summarizes the analytical populations used for each component of the study. Panel B shows the four mutually exclusive clinical phenotypes among patients in whom both ocular and palatal status were explicitly documented. OPT, oculopalatal tremor; HOD, hypertrophic olivary degeneration.

| **A. Analytical populations** |  |  |
|---|---|---|
| **Analytical population** | **n** | **Primary purpose** |
| All patient records | 501 | Demographics, lesion and HOD distribution |
| Ocular status explicitly documented | 230 | Predictors of ocular oscillations |
| Palatal status explicitly documented | 315 | Predictors of palatal oscillations |
| Both ocular and palatal status documented | 187 | Four-level phenotype classification |
| Complete phenotype + lesion site + etiology + HOD side | 142 | Phenotype-specific predictor analyses |
| Complete phenotype + onset interval | 45 | Phenotype timing |
| OPT + explicit onset interval | 35 | Time to OPT development |
| **B. Complete clinical phenotype** |  |  |
| **Phenotype** | **n** | **%** |
| Oculopalatal tremor | 118 | 63.1 |
| Isolated ocular oscillation | 21 | 11.2 |
| Isolated palatal oscillation | 8 | 4.3 |
| Neither | 40 | 21.4 |
| Total | 187 | 100.0 |

Among 187 patients in whom both ocular and palatal manifestations were definitively documented, 118 (63.1%) had OPT, 21 (11.2%) had isolated ocular oscillations, 8 (4.3%) had isolated palatal oscillations, and 40 (21.4%) had neither manifestation (Table 1). The remaining 314 patients could not be assigned reliably because one or both manifestations were not explicitly documented.

### Hypertrophic Olivary Degeneration (HOD)

#### HOD Frequency and Laterality

HOD status was definitively reported in 460 patients. HOD was present in 450/460 (97.8%) and absent in 10/460 (2.2%). This high frequency reflects selection of the dataset around HOD and related syndromes and prevents meaningful inference regarding predictors. HOD laterality was classifiable in 424 patients. HOD was unilateral in 221/424 (52.1%) and bilateral in 203/424 (47.9%). Among unilateral cases with documented side, 113/212 (53.3%) were left-sided and 99/212 (46.7%) were right-sided, with no meaningful overall left-right predominance.

#### Primary Lesion Location Predicts HOD Laterality

Primary lesion location was strongly associated with unilateral versus bilateral HOD (chi-square=35.65, p=3.4 x 10^-7; Cramer V=0.31). Focal brainstem or cerebellar lesions predominantly produced unilateral HOD, whereas combined pathway involving multiple posterior fossa structures, and other anatomically extensive lesions were more frequently associated with bilateral HOD.

#### Lesion Side Predicts the Side of HOD

The relationship between lesion side and HOD became anatomically coherent after stratification by lesion level. Among 70 classifiable lateralized brainstem/pons lesions, 49 (70.0%) produced ipsilateral HOD, only 3 (4.3%) produced isolated contralateral HOD, and 18 (25.7%) produced bilateral HOD (Table 2). Conversely, among 44 lateralized cerebellar lesions, 38 (86.4%) produced contralateral HOD, 5 (11.4%) produced bilateral HOD, and only 1 (2.3%) produced isolated ipsilateral HOD. Combined pathway lesions were relatively more likely to produce bilateral HOD. This lesion-level relationship was one of the strongest anatomically coherent findings in the dataset (Table 2).

**Table 2.**
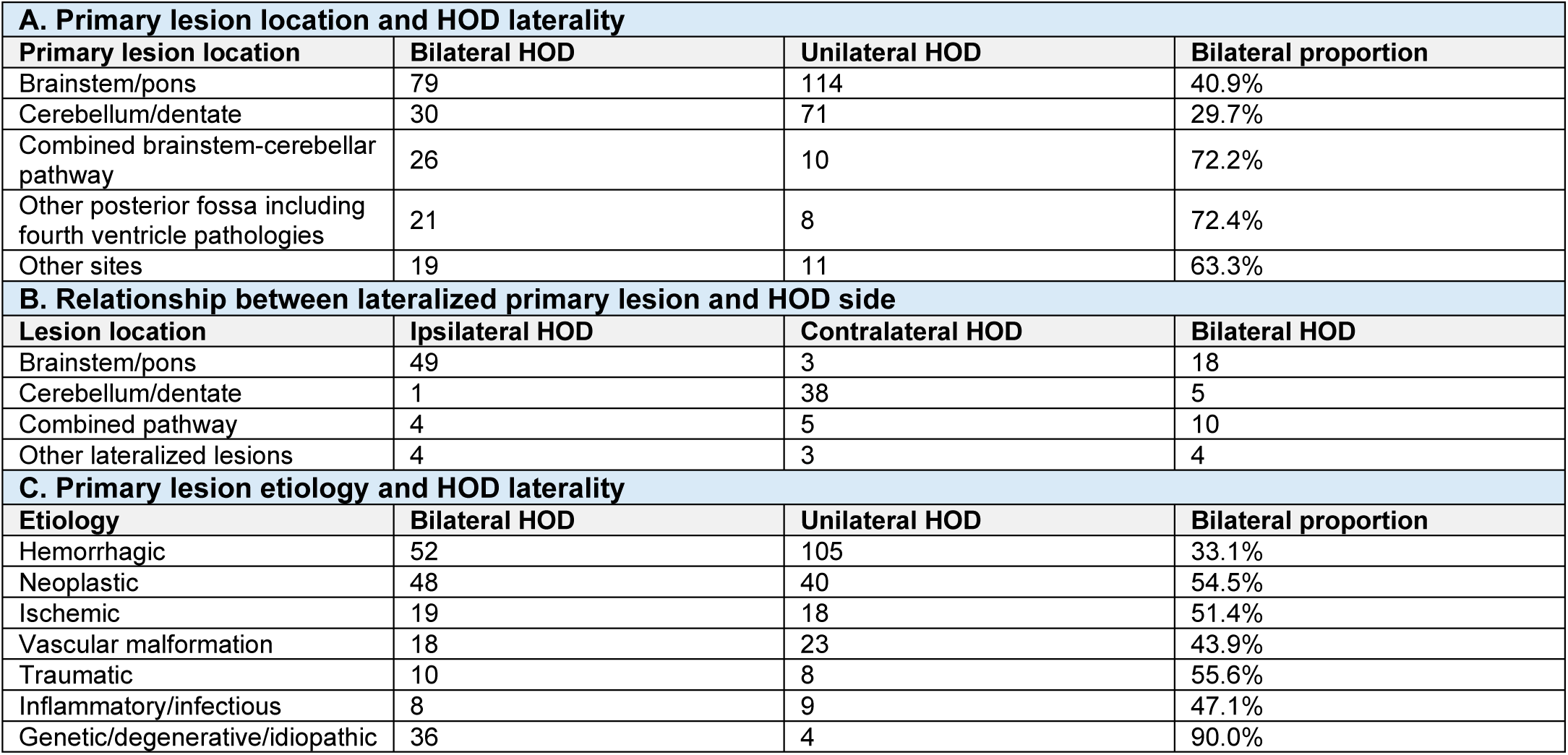
Relationship of primary lesion characteristics to HOD laterality and side. Panel A shows the association between primary lesion location and unilateral versus bilateral HOD. Panel B shows the side of HOD relative to a clearly lateralized primary lesion; ipsilateral and contralateral refer to the relationship between the primary lesion and the affected inferior olive. Panel C shows HOD laterality across etiologic groups. The posterior fossa/fourth ventricle category consisted predominantly of anatomically complex posterior-fossa and fourth-ventricular lesions, often tumor-related and postoperative. HOD, hypertrophic olivary degeneration.

#### Primary Lesion Etiology and HOD Laterality

Etiology was also associated with unilateral versus bilateral HOD (chi-square=27.35, p=0.0012; Cramer V=0.27; Table 2). Hemorrhagic lesions most commonly produced unilateral HOD. Neoplastic, ischemic, traumatic, and inflammatory lesions showed more balanced unilateral-bilateral distributions. Genetic, degenerative, and idiopathic disorders were overwhelmingly bilateral, likely reflecting diffuse or bilateral pathway involvement. Etiology and anatomical extent were not independent, and this association should therefore not be interpreted as a direct biological effect of lesion cause.

### Predictors of Any Ocular Oscillations

Ocular examination was explicitly documented in 230 patients. Ocular oscillations were present in 182/230 (79.1%). Among patients with classifiable HOD laterality, ocular oscillations occurred in 70/84 (83.3%) with bilateral HOD compared with 62/95 (65.3%) with unilateral HOD. Bilateral HOD was therefore associated with substantially greater odds of ocular oscillations (OR 2.66, 95% CI 1.31-5.43; Fisher exact p=0.0067).

Primary lesion location was associated with ocular oscillations (omnibus p=0.0079; Cramer V=0.26). Ocular oscillations occurred in 12/14 (85.7%) patients with combined pathway lesions, 110/135 (81.5%) with brainstem/pons lesions, 25/37 (67.6%) with cerebellar/dentate lesions, 8/12 (66.7%) with other sites, and 1/5 (20.0%) with posterior fossa/fourth ventricular lesions. Brainstem and combined pathway lesions therefore showed the highest frequency of ocular manifestations.

Etiology was also associated with ocular oscillations (omnibus p=0.0050; Cramer V=0.30). Because several etiologic strata were small, this result was interpreted primarily through the larger hemorrhagic, ischemic, and neoplastic groups rather than through extreme proportions in small, selected categories.

### Predictors of Any Palatal Oscillations

Palatal examination was explicitly documented in 315 patients. Palatal oscillations were present in 197/315 (62.5%). Among patients with classifiable HOD laterality, palatal oscillations occurred in 80/125 (64.0%) with bilateral HOD and 79/145 (54.5%) with unilateral HOD. The association was weaker than that observed for ocular oscillations (OR 1.49, 95% CI 0.91-2.42; Fisher exact p=0.137). Primary lesion location was not significantly associated with palatal oscillations (p=0.250). Etiology showed a nominal association (p=0.031; Cramer V=0.24), but interpretation was limited by sparse categories.

The preceding ocular analysis compared patients with versus without any ocular oscillation, irrespective of whether palatal oscillations were also present; and vice versa for palatal analysis irrespective of ocular involvement. On the contrary complete-phenotype analysis addressed a different question and was restricted to patients in whom both ocular and palatal status were explicitly documented and classified each patient into combined OPT, ocular-only, palatal-only, or neither. This analysis determined whether lesion location, etiology, or HOD pattern distinguished the specific clinical phenotype, rather than simply predicting the presence of an ocular or palatal manifestation.

### Predictors of the Complete Clinical Phenotype

The complete predictor cohort comprised 142 patients: 77 with OPT, 19 with ocular-only manifestations, 8 with palatal-only manifestations, and 38 with neither phenotype (Table 3).

**Table 3.** Clinical phenotype according to HOD pattern, primary lesion location, and lesion etiology. The association cohort included 142 patients with complete ocular and palatal phenotype classification, primary lesion location, lesion etiology, and HOD side. Panel A shows phenotype according to HOD pattern; Panel B according to primary lesion location; and Panel C according to primary lesion etiology. The four phenotype categories are mutually exclusive: OPT indicates both ocular and palatal oscillations; ocular only indicates ocular oscillations with palatal oscillations absent; palatal only indicates palatal oscillations with ocular oscillations absent; neither indicates both manifestations absent. OPT, oculopalatal tremor; HOD, hypertrophic olivary degeneration.

| **A. HOD pattern according to clinical phenotype** |  |  |  |  |
| **HOD pattern** | **OPT** | **Ocular only** | **Palatal only** | **Neither** |
|---|---|---|---|---|
| Bilateral | 37 | 11 | 5 | 9 |
| Unilateral left | 23 | 5 | 1 | 15 |
| Unilateral right | 17 | 3 | 2 | 14 |
| **B. Clinical phenotype by primary lesion location** |  |  |  |  |
| **Lesion location** | **OPT** | **Ocular only** | **Palatal only** | **Neither** |
| Brainstem/pons | 57 | 13 | 2 | 22 |
| Cerebellum | 9 | 4 | 2 | 10 |
| Combined pathway | 8 | 1 | 0 | 2 |
| Other sites | 3 | 1 | 3 | 1 |
| Posterior fossa/fourth ventricle | 0 | 0 | 1 | 3 |
| **C. Clinical phenotype by primary lesion etiology** |  |  |  |  |
| **Etiology** | **OPT** | **Ocular only** | **Palatal only** | **Neither** |
| Hemorrhagic | 50 | 8 | 2 | 24 |
| Neoplastic | 7 | 5 | 5 | 9 |
| Ischemic | 6 | 4 | 0 | 1 |
| Vascular malformation | 7 | 0 | 0 | 3 |
| Inflammatory/infectious | 5 | 2 | 0 | 0 |
| Other | 2 | 0 | 1 | 1 |

#### HOD pattern

When left and right unilateral HOD were combined, bilateral HOD was associated with OPT versus neither (OR 2.98, 95% CI 1.25-7.12; p=0.015), ocular only versus neither (OR 4.43, 95% CI 1.36-14.40; p=0.018), and palatal only versus neither (OR 5.37, 95% CI 1.07-27.00; p=0.044). The palatal-only estimate was based on only eight events and was therefore highly unstable. When unilateral left, unilateral right, and bilateral HOD were retained as three separate categories, the overall four-phenotype association did not reach significance (p=0.114; Cramer V=0.19). The data therefore provided no convincing evidence that left and right unilateral HOD generate different phenotypes; the principal distinction was bilateral versus unilateral involvement.

#### Primary Lesion Location and Phenotype

Primary lesion location significantly differentiated the four phenotypes (chi-square=32.78, p=0.0010; Cramer V=0.28; Table 3). Combined pathway lesions had the highest proportion of OPT (72.7%), followed by brainstem/pons lesions (60.6%) and cerebellar lesions (36.0%). Posterior fossa/fourth ventricular lesions rarely produced the complete phenotype in this small subgroup.

When brainstem/pons lesions were compared with cerebellar lesions for OPT versus neither phenotype, the OR was 2.88 (95% CI 1.03-8.03), with a borderline Fisher exact p value of 0.056. The magnitude and direction of the effect, together with the significant omnibus association, supported an anatomical tendency favoring OPT after brainstem rather than isolated cerebellar lesions.

### Etiology and Clinical Phenotype

Etiology differed across the four phenotypes (chi-square=34.54, p=0.0108; Cramer V=0.28). Because several expected counts were small, this omnibus association was considered exploratory (Table 3).

#### Hemorrhagic lesions

Among hemorrhagic cases, 59.5% had OPT, 9.5% isolated ocular oscillations, 2.4% isolated palatal oscillations, and 28.6% neither manifestation. Thus, when an oscillatory phenotype followed hemorrhage, it was most commonly the combined OPT phenotype.

#### Ischemic lesions

Among ischemic cases, 54.5% had OPT and 36.4% isolated ocular oscillations; none had isolated palatal oscillations. Compared with hemorrhage, ischemia was associated with substantially greater odds of ocular-only disease relative to the “neither” phenotype (OR 12.0, 95% CI 1.16-123.69; p=0.030). The wide confidence interval reflected the small ischemic subgroup.

#### Neoplastic lesions

Neoplastic lesions produced the most heterogeneous phenotype: 26.9% OPT, 19.2% ocular only, 19.2% palatal only, and 34.6% neither. Compared with hemorrhage, neoplastic lesions showed directionally lower odds of OPT versus neither (OR 0.37; p=0.090). Neoplasia was disproportionately represented among isolated palatal cases (palatal only vs neither, neoplastic vs hemorrhagic: OR 6.67, 95% CI 1.09-40.74; p=0.039). This association may reflect lesion extent, progressive posterior fossa distortion, surgery, radiation, recurrence, or involvement of multiple pathway components rather than tumor biology itself.

#### Vascular malformations

Among nonhemorrhagic vascular-malformation cases, 70% had OPT and 30% had neither manifestation; no completely classified ocular-only or palatal-only cases were identified. Overall, the etiologic findings suggested that hemorrhagic and vascular-malformation lesions preferentially produced combined OPT when an oscillatory phenotype occurred, ischemic lesions showed relatively greater representation of isolated ocular manifestations, and neoplastic/post-treatment lesions produced a more heterogeneous phenotype.

### Predictive Modeling Among Patients with Confirmed HOD

The confirmed-HOD predictive cohort included 159 patients: 91 with OPT, 20 with ocular-only oscillations, 8 with palatal-only oscillations, and 40 with neither manifestation. This analysis treated the four mutually exclusive phenotypes as the ground-truth outcome and used only demographic and lesion characteristics that could precede the clinical phenotype. Etiology differed across outcomes (p=0.0024). Vascular malformations and other structural causes were enriched among OPT cases, ischemic lesions showed relatively greater representation of ocular-only disease, and neoplastic lesions were disproportionately represented among palatal-only cases. Lesion location showed an anatomical trend (p=0.082): OPT occurred in 75.0% of combined pathway lesions and 60.8% of brainstem/pons lesions, compared with 36.0% of cerebellar/dentate lesions. Age (p=0.556), sex (p=0.219), and primary-lesion laterality (p=0.746) did not differentiate phenotype.

The antecedent-factor model included age, sex, lesion etiology, lesion location, and lesion laterality. Cross-validated four-class discrimination was limited: macro AUC was 0.555 (SD 0.077), balanced accuracy 0.301 (SD 0.091), macro F1 score 0.262 (SD 0.070), and log loss 1.370 (SD 0.083) (Table 4). For comparison, chance-level balanced accuracy for four classes was 0.25. The strongest directional model coefficients were vascular-malformation etiology for OPT, ischemic etiology for ocular-only disease, and neoplastic etiology and atypical lesion location for palatal-only disease; these coefficients were shrinkage estimates and were not interpreted as conventional inferential odds ratios.

**Table 4.**
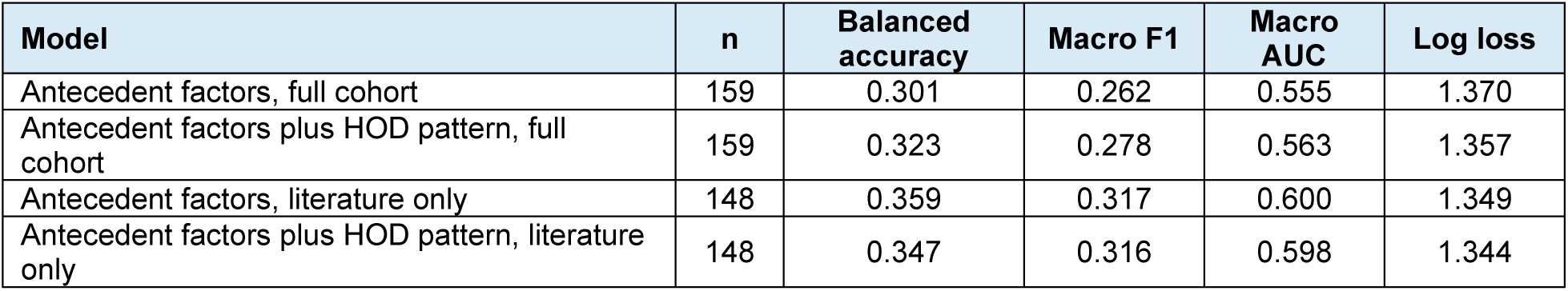
Cross-validated performance of multinomial phenotype prediction models among patients with confirmed HOD. The primary model included age, sex, primary-lesion etiology, lesion location, and lesion laterality. The imaging-enhanced model additionally included HOD laterality. Literature-only models excluded the clinically referred Cleveland cases. Values are means across repeated stratified four-fold cross-validation. Balanced accuracy and macro F1 weight each phenotype equally. AUC, area under the receiver operating characteristic curve; HOD, hypertrophic olivary degeneration.

| **Model** | **n** | **Balanced accuracy** | **Macro F1** | **Macro AUC** | **Log loss** |
|---|---|---|---|---|---|
| Antecedent factors, full cohort | 159 | 0.301 | 0.262 | 0.555 | 1.370 |
| Antecedent factors plus HOD pattern, full cohort | 159 | 0.323 | 0.278 | 0.563 | 1.357 |
| Antecedent factors, literature only | 148 | 0.359 | 0.317 | 0.600 | 1.349 |
| Antecedent factors plus HOD pattern, literature only | 148 | 0.347 | 0.316 | 0.598 | 1.344 |

Adding HOD laterality produced only a small improvement in discrimination: macro AUC increased to 0.563 (SD 0.076), balanced accuracy to 0.323 (SD 0.083), and macro F1 to 0.278 (SD 0.058), while log loss decreased to 1.357 (SD 0.079). Bilateral HOD was directionally associated with an oscillatory phenotype, whereas right-versus-left unilateral HOD added little information. In a literature-only sensitivity analysis excluding the clinically referred Cleveland cases, macro AUC was 0.600 for the antecedent model and 0.598 after HOD laterality was added, confirming that the imaging variable did not materially improve discrimination (Table 4). A hierarchical analysis did not resolve the limited prediction. For any ocular or palatal oscillation versus neither manifestation, the antecedent model yielded an AUC of 0.508 and the imaging-enhanced model an AUC of 0.556. Among symptomatic patients, discrimination of OPT from ocular-only and palatal-only phenotypes remained modest (macro AUC 0.570 without and 0.557 with HOD laterality). These results indicate that the measured factors define population-level associations but do not provide sufficient separation for an individual-level clinical prediction tool.

### Ocular Phenomenology

#### Oculopalatal Tremor

Among 118 patients with OPT, ocular waveform was classifiable in 65. Fifty-nine had a pendular waveform, 2 had a pendular waveform with a jerk component, 2 had other explicitly pendular variants, 1 had a pure jerk waveform, and 1 had rapid jerky oscillations. The described directions included vertical, vertical-torsional, torsional, multiplanar horizontal-vertical-torsional, and dissociated or asymmetric vertical oscillations.

#### Isolated Ocular Oscillations

Among 21 ocular-only patients, six had explicitly pendular oscillations and one had an irregular rhythmic waveform; waveform was not reported in 14. Reported manifestations were often heterogeneous and included pendular, gaze-evoked, upbeat, horizontal, mixed vertical-horizontal, and dissociated nystagmus.

### Palatal Phenomenology

Terminology used for palatal manifestations included palatal tremor, palatal myoclonus, symptomatic palatal tremor, palatal/pharyngeal tremor, and palatopharyngeal or palatolaryngeal myoclonus. In these reports, use of “tremor” versus “myoclonus” appeared to reflect nomenclature rather than demonstrably different physiology.

### Associated Neurological and Movement-Disorder Phenotypes

Associated neurological manifestations were explicitly documented in 243 patients. Ataxia was the dominant associated manifestation. Among OPT patients with sufficiently documented associated findings, 52/73 (71.2%) had ataxia, 30/73 (41.1%) weakness or plegia, 19/73 (26.0%) another tremor, and 10/73 (13.7%) dystonia. Among ocular-only patients with documented associated findings, 13/16 (81.3%) had ataxia. All four palatal-only patients with documented associated manifestations had ataxia, although this denominator was too small for comparative inference. Chorea was exceptionally uncommon.

### Latency From Primary Lesion to Clinical Manifestation

#### Overall Timing

An explicit symptom-onset interval was reported in 105 patients. Across these patients, median onset was 7 months (IQR 4-17 months), mean onset was 19.2 months, and the range extended from immediate onset to 288 months. The mean was strongly influenced by a small number of very delayed cases; medians and IQRs were therefore considered more representative. Among 45 patients in the complete-phenotype cohort with timing information, median onset was 7 months (IQR 4.5-18 months).

#### OPT-Specific Timing

Timing was available in 35 patients with OPT. Median onset was 6 months (IQR 4.75-24 months), mean onset was 29.3 months, and the range extended from immediate onset to 288 months.

Cumulative onset was 5.7% by 1 month, 20.0% by 3 months, 51.4% by 6 months, 65.7% by 12 months, and 77.1% by 24 months. Thus, approximately half of documented OPT cases became clinically apparent by six months, approximately two thirds by one year, and more than three quarters by two years. Delayed onset several years after the causative lesion was documented but uncommon.

#### Do Anatomical or Etiologic Variables Predict OPT Latency?

No convincing predictor of onset latency emerged. There were no statistically significant differences according to etiology (p=0.267), lesion location (p=0.527), or HOD laterality/side (p=0.841).

MRI was performed or HOD was identified at a median of approximately 6 months (IQR 3-12 months), but MRI timing was determined by clinical practice and should not be interpreted as biological onset of HOD.

#### Do demographics predict ocular or palatal oscillations?

Age was available in 456 patients. Median age was 51 years (IQR 36.8-60.3; range 1.5-89 years). There were 280 male and 172 female patients; sex was not reported in 49. Neither age nor sex predicted ocular or palatal oscillations (age and ocular oscillations, p=0.364; age and palatal oscillations, p=0.268; sex and ocular oscillations, p=0.489; sex and palatal oscillations, p=0.904). Anatomical characteristics were therefore substantially more informative than basic demographic variables.

## Discussion

This pooled patient-level analysis provides a large-scale examination of the anatomical, etiologic, and clinical determinants of OPT and related manifestations of HOD. Several findings emerged consistently across complementary analyses. First, the anatomical level of the causative lesion strongly determined the laterality of subsequent HOD: brainstem lesions produced predominantly ipsilateral HOD, whereas cerebellar (deep cerebellar nuclei) lesions produced contralateral HOD. Second, bilateral HOD was substantially more strongly associated with ocular oscillations than with palatal oscillations. Third, the complete OPT phenotype was preferentially associated with brainstem and combined pathway lesions, while isolated cerebellar/dentate lesions were less likely to produce combined OPT. Fourth, right-versus-left unilateral HOD had little apparent relationship to phenotype; bilaterality was more informative than side. Fifth, classical OPT was overwhelmingly characterized by “pendular” ocular oscillations. Finally, OPT typically became clinically apparent several months after the inciting lesion, with median onset around six months, but the available anatomical variables did not reliably predict either phenotype or latency in an individual patient.

Together, these observations support the concept that HOD represents a substrate for abnormal oscillation but does not itself determine the clinical syndrome. The anatomical extent of network injury and subsequent olivocerebellar reorganization appear to be critical determinants of whether HOD is expressed as ocular tremor, palatal tremor, combined OPT, or no clinically apparent oscillation.

Lesion level accounting for the laterality of HOD is an intuitive result. More focal lesions affecting brainstem, pons, or cerebellum leads to focused disruption of descending central tegmental tract fibers leading to unilateral HOD. Central tegmental track does not cross, leading to ipsilateral deficit, while interruption of cerebellar outflow that cross in the superior cerebellar peduncle causes contralateral deficit. Combined pathway lesions, being more extensive, produce bilateral HOD.

Inferior olivary deafferentation has been implicated in symptomatic palatal tremor^3^. The oscillator-amplifier model proposes that synchronized olivary activity provides a low-frequency signal that maladaptive cerebellar learning amplifies and reshapes into complex ocular oscillations^14, 15, 19–21^. Bilateral HOD may mark more extensive involvement of this network, increasing the likelihood that pathological activity is expressed in ocular motor circuits, favoring ocular oscillations by engaging more of the olivocerebellar network. Consistent with this possibility, ocular oscillations were more frequent with bilateral than unilateral HOD (83.3% vs 65.3%, OR 2.66). Among fully characterized cases, bilateral HOD was associated with approximately threefold higher odds of combined OPT and more than fourfold higher odds of ocular-only oscillations, each relative to neither manifestation. The same concept also explains why combined pathway lesions had the highest proportion of OPT, followed by brainstem lesions.

A pendular component was present in 96.9% of OPT cases with documented waveform, often with vertical, torsional, or mixed components and interocular asymmetry^11, 14, 20–22^. The ocular-only group also included gaze-evoked, upbeat, and horizontal nystagmus, some of which may reflect the primary lesion itself. It is noteworthy that number of cases included in this analysis offered clinical characterization. On the contrary, objective measure of oculographic waveform highlights non-pendular, coarse irregular eye oscillations; these are neither jerk nor pendular; but represent combined effects of more than one oscillators as well as mal-adaptive cerebellar learning^14, 21, 22^. Among OPT cases with documented associated features, more than 70% had ataxia and approximately 14% had dystonia. The frequency of ataxia places OPT within a broader cerebellar and brainstem syndrome, as well as maladaptive state induced by olive-cerebellar hypersynchrony suppressing normal motor learning^14, 19, 20, 23^. Dystonia is becoming increasingly recognized feature accompanying feature of the disease and it could represent manifestation of aberrant cerebellar hyperactivity transmitted to the basal ganglia, making the circuit unstable^14, 19, 20^.

Hemorrhage tended to accompany combined OPT and similar pattern was seen with vascular malformation. This is an expected finding since vascular malformation often leads to hemorrhage in corresponding location. Ischemia more often accompanied ocular-only oscillations, and neoplasia was overrepresented among isolated palatal cases. Ischemia and tumor both are likely to be focal, presenting more limited involvement, and isolated presentation. In other words, these exploratory patterns may reflect differences in lesion location, extent, chronicity, or treatment, including surgery and radiation.

The timing analysis provides a clinically useful characterization of OPT evolution. Among 35 OPT patients with explicit timing, median onset was approximately six months. About half had developed OPT by six months, approximately two thirds by one year, and more than three quarters by two years.

This delayed clinical course corresponds broadly to the known temporal evolution of HOD. Increased olivary T2 signal may appear approximately one month after pathway injury, while frank hypertrophy generally emerges later, around six months^10^. The similarity between the approximate timing of olivary hypertrophy and median OPT onset is biologically appealing, but the two measures should not be conflated. MRI timing in this dataset reflects when imaging was obtained, not when olivary pathology first developed.

One of the most important findings was the discrepancy between biologically coherent group-level associations and poor individual-level predictive performance. In the confirmed-HOD cohort, lesion etiology differentiated phenotype, and bilateral HOD was more frequent among patients with ocular manifestations. Nevertheless, the antecedent four-class model achieved a macro AUC of only 0.555 and balanced accuracy of 0.301; adding HOD laterality increased these values only to 0.563 and 0.323. Hierarchical modeling of any oscillatory phenotype versus neither also remained near chance. This is not contradictory. An odds ratio or group association indicates that a feature is more common in one phenotype than another; it does not imply sufficient sensitivity, specificity, or independence to identify an individual patient’s future phenotype. Considerable anatomical overlap remained among OPT, ocular-only, palatal-only, and clinically unaffected patients. Moreover, the available predictors were coarse structural descriptors and did not capture exact tract-level lesion topography, inferior olivary subnuclei, quantitative hypertrophy, residual pathway connectivity, cerebellar adaptive gain, ocular motor integrator involvement, or longitudinal network evolution. The present analyses should therefore be regarded as predictor-association models rather than clinical prediction tools. Future predictive studies will require quantitative lesion mapping, tractography, longitudinal imaging, quantitative eye-movement recordings, and functional measures of cerebellar-olivary connectivity.

The data support several clinically useful principles. First, the side of HOD should be interpreted according to the anatomical level of the primary lesion. Brainstem lesions generally produce ipsilateral HOD, whereas cerebellar/dentate lesions generally produce contralateral HOD. Second, bilateral HOD increases the probability of an ocular manifestation and of combined OPT. Bilaterality is more clinically informative than whether unilateral HOD is right- or left-sided. Third, patients with combined brainstem-cerebellar pathway injury appear particularly likely to develop the complete OPT syndrome, whereas isolated cerebellar/dentate lesions can produce HOD without necessarily producing OPT. Fourth, OPT typically emerges over months rather than immediately after the lesion. Clinical surveillance should therefore extend beyond the acute neurological event, particularly through the first one to two years. These associations can inform clinicoradiological interpretation and patient counseling but are not sufficiently discriminative to predict an individual patient’s outcome with confidence.

This study has several limitations. Ocular and palatal status were documented in 230 and 315 of 501 patients, respectively, and four-class phenotype assignment required explicit documentation of both findings. Most cases came from retrospective reports or series with variable examination and imaging protocols. Publication and referral bias, shared study-level ascertainment, and broad lesion categories limit generalizability. Primary-lesion laterality was unavailable in more than half of the confirmed-HOD predictive cohort. Only eight isolated palatal cases and 35 timed OPT cases were available, limiting phenotype-specific and latency analyses. Multiple exploratory comparisons further limit inference, particularly for small etiologic and phenotypic subgroups.

In conclusion, this pooled patient-level analysis demonstrates that the clinical expression of hypertrophic olivary degeneration is determined more strongly by the location and extent of cerebello-rubro-olivary pathway disruption than by demographic characteristics or right-versus-left olivary involvement. The side of HOD follows the known anatomy of the pathway: lateralized brainstem lesions predominantly produce ipsilateral HOD, whereas cerebellar/dentate lesions predominantly produce contralateral HOD. Bilateral HOD preferentially associates with ocular oscillations and increases the likelihood of the combined OPT phenotype, while the relationship with palatal oscillations alone is substantially weaker. Combined brainstem-cerebellar pathway lesions show the greatest propensity for complete OPT, whereas isolated cerebellar/dentate lesions are less likely to produce the full syndrome. Classical OPT is overwhelmingly characterized by pendular ocular oscillations, most often involving vertical, torsional, or mixed planes. OPT generally develops after a delay, with a median onset of approximately six months and more than three quarters of documented cases emerging within two years. These findings support a distributed network model in which inferior olivary degeneration creates the substrate for pathological oscillation but does not alone determine clinical expression. The extent of pathway injury, bilateral olivary involvement, and subsequent cerebellar network adaptation likely determine whether the resulting syndrome manifests as ocular tremor, palatal tremor, combined OPT, or no overt oscillatory phenotype. Although these anatomical and etiologic variables provide meaningful population-level associations, their limited cross-validated discrimination indicates that they cannot yet reliably predict the phenotype of an individual patient.

## Data Availability

Data will be shared with interested parties after appropriate institutional agreements are put in place.

## Author’s Roles

- HA: data collection, analysis, review and edit draft
- IC: data collection, analysis, review and edit draft
- AG: conceptualize idea, data collection, analysis, write and edit draft

## Supplementary Methods

### S1 Supplementary References

https://pubmed.ncbi.nlm.nih.gov/?term=longquery39fc0afc0405e66a5a45&sort=date

### S2 Literature Search and Case Ascertainment

The pooled patient-level dataset consisted of 469 cases from a previously assembled literature extraction and 32 previously unpublished cases from our clinical practice. The patient was the unit of analysis. We treated the pooled records as a heterogeneous observational dataset because source reports varied in referral populations, clinical assessments, imaging protocols, terminology, and completeness of reporting. We pooled the clinical series with the literature cases for the principal clinicoradiologic analyses and examined the clinical series separately to assess potential referral effects. The University Hospitals Cleveland Medical Center Institutional Review Board approved the retrospective chart review of the unpublished clinical cases.

We conducted a structured PubMed search from database inception through June 2026. Search terms included “oculopalatal tremor,” “oculopalatal myoclonus,” “inferior olive hypertrophy,” “hypertrophic inferior olivary degeneration,” and “progressive ataxia palatal tremor.” We screened reports for patients with hypertrophic olivary degeneration (HOD), ocular or palatal oscillations associated with lesions affecting the cerebello-rubro-olivary (Guillain–Mollaret) pathway, or both. We reviewed relevant articles for extractable clinical, anatomical, imaging, etiologic, phenomenologic, and temporal data at the individual-patient level. When publications from the same author group described patients with similar clinical characteristics, lesion etiologies, imaging findings, or other identifying features, we compared the reports for potential overlap. We included patients judged to have been reported more than once only once in the pooled dataset.

### S3 Data Abstraction and Missing Data

Using a standardized approach, we recorded age and sex, primary lesion location and side, primary lesion etiology, HOD presence and laterality, ocular and palatal oscillation status, ocular waveform and direction when reported, associated neurologic and movement-disorder manifestations, time from the causative lesion to symptom onset, timing of MRI demonstrating or evaluating HOD, and treatment information when available.

Two reviewers independently evaluated a randomly selected 30% of included publications. We compared independent assessments for agreement on study eligibility, patient identification, and clinical and imaging variables. The reviewers resolved discrepancies by consensus. We did not calculate a formal interrater reliability statistic. Analyses used the records with sufficient patient-level information for the variables under study. Denominators therefore varied by analysis. Unreported ocular, palatal, associated neurologic, treatment, and timing data were coded as missing.

### S4 Primary Lesion Location and Etiology

We grouped lesion locations as brainstem or pons, cerebellum or dentate nucleus, combined brainstem– cerebellar or multicomponent pathway involvement, posterior fossa or fourth ventricle, and other sites. We grouped etiologies as hemorrhagic, ischemic, neoplastic, vascular malformation, traumatic, inflammatory or infectious, genetic or degenerative or idiopathic, and other causes. When hemorrhage associated with a vascular malformation was classified as the primary lesion, we assigned the case to the hemorrhagic category alone.

### S5 Definition and Laterality of HOD

HOD status followed the radiographic interpretation in the source record. We classified HOD as unilateral or bilateral and recorded left or right for unilateral cases when the side was available. Analyses relating lesion side to HOD included only patients with a clearly lateralized primary lesion and classifiable HOD. We coded HOD as ipsilateral, contralateral, or bilateral relative to the causative lesion.

### S6 Clinical Phenotype and Analysis Cohorts

We assigned one of four mutually exclusive phenotypes only when the source explicitly documented both ocular and palatal oscillation status. Combined oculopalatal tremor (OPT) required both to be present. The ocular-only phenotype required ocular oscillations to be present and palatal oscillations to be absent. The palatal-only phenotype required palatal oscillations to be present and ocular oscillations to be absent. The neither phenotype required both to be absent.

The complete phenotype cohort included 187 patients. Association analyses requiring classifiable HOD side or laterality, primary lesion location, and primary lesion etiology included 142 patients. For predictive modeling, we used the 159 patients with confirmed HOD and complete four-class phenotype assignment. This predictive cohort included 91 patients with OPT, 20 with ocular-only oscillations, 8 with palatal-only oscillations, and 40 with neither manifestation.

### S7 Ocular and Palatal Phenomenology

We classified ocular waveform and direction using the terminology in each source report. We coded pendular, pendular-with-jerk, and other explicitly pendular waveforms as having a pendular component. We retained separate descriptions for gaze-evoked, upbeat, horizontal, mixed, dissociated, and other nonpendular patterns. We retained the reported palatal terminology because historical use of “tremor” and “myoclonus” did not consistently distinguish underlying physiology. Phenomenologic analyses were primarily descriptive given incomplete waveform documentation.

### S8 Associated Neurologic Manifestations

We grouped associated manifestations as ataxia, weakness or plegia, tremor other than palatal tremor, sensory abnormalities, dystonia, and chorea. Percentages used the denominator of patients with explicit documentation of associated neurologic manifestations.

### S9 Symptom and MRI Timing

We extracted the interval from the primary lesion to clinical symptom onset when explicitly reported. We analyzed symptom timing among all patients with a documented onset interval, within the complete phenotype cohort, and within the OPT subgroup. We analyzed MRI timing separately. The interval from the causative lesion to an MRI demonstrating or evaluating HOD represented the timing of imaging observation. The date of first biological onset of HOD could not be inferred from this interval.

### S10 Statistical Analysis

We summarized categorical variables using counts and percentages and continuous variables primarily using medians and interquartile ranges because several distributions, particularly symptom-onset latency, were markedly right-skewed. We used Pearson chi-square tests for multigroup categorical comparisons and calculated Cramér V to describe association strength. We used Fisher exact tests for two-group comparisons and when individual categories were sparse.

We first examined associations of lesion location, lesion etiology, and HOD pattern with the four-level phenotype using omnibus tests. We then performed focused pairwise comparisons for anatomically or clinically relevant contrasts, reporting odds ratios and 95% confidence intervals. The group with neither ocular nor palatal oscillations was the primary reference for comparisons with OPT, ocular-only, and palatal-only phenotypes.

Because some outcomes were uncommon, particularly isolated palatal oscillations, we used class-balanced L2-penalized multinomial logistic regression. The primary antecedent-factor model included age, sex, primary-lesion etiology, primary-lesion location, and primary-lesion laterality. Etiology was consolidated as hemorrhagic, ischemic, neoplastic, vascular malformation, or other/unclear. Lesion location was consolidated as brainstem/pons, cerebellum/dentate, combined brainstem-cerebellar pathway, or other/unclear. Lesion laterality was classified as left, right, bilateral/midline, or unknown. Age was standardized; categorical variables were dummy coded within each training fold. A secondary imaging-enhanced model added HOD pattern as bilateral, left unilateral, right unilateral, or unavailable.

Models used an L2 penalty with inverse regularization strength C=0.5 and balanced class weights. We estimated out-of-sample performance with repeated stratified four-fold cross-validation using five repetitions for the principal models. Performance measures were overall accuracy, balanced accuracy, macro F1 score, macro one-versus-rest AUC, and multiclass log loss. We emphasized balanced accuracy, macro F1, and macro AUC because the four outcomes were imbalanced. Coefficients from the penalized full-cohort model were interpreted directionally as shrinkage estimates, not as conventional inferential odds ratios.

We performed two sensitivity analyses. First, we repeated the primary and imaging-enhanced models after excluding the clinically referred Cleveland cases. Second, we used a hierarchical approach that compared any oscillatory phenotype with neither manifestation and then, among symptomatic patients, distinguished OPT, ocular-only, and palatal-only phenotypes. These analyses tested whether collapsing the outcome into clinically simpler stages improved discrimination. Analyses were performed in Python 3.12 using scikit-learn 1.8.0 and SciPy 1.17.0.

We compared onset latency across lesion etiology, lesion location, and HOD laterality groups with Kruskal– Wallis tests. All statistical tests were two-sided. Given the exploratory design and correlated comparisons, we interpreted p values in the context of effect sizes, confidence intervals, sample sizes, biological plausibility, and consistency across related analyses.

